# The Pediatric Outcomes Data Collection Instrument (PODCI) as Performance Measure, Comparing General Population with Cerebral Palsy Population Using the Gross Motor Function Classification System Levels I-V

**DOI:** 10.64898/2026.04.01.26349726

**Authors:** Chauna N. Weyermuller, Joseph Andary, Demiana Soliman, Philip E. Gates

## Abstract

**OBJECTIVES:** Compare results of the Pediatric Outcomes Data Collection Instrument (PODCI) in children ages 2-18 years with cerebral palsy (CP) across all severity levels of the Gross Motor Function Classification System (GMFCS) with children in the General Population, confirming discriminant validity as a performance assessment tool and health-related quality of life (HRQOL) measure.

**METHODS:** Cross-sectional study: single response PODCI proxy survey databases of 5238 children ages 2-18 years in GP and 2470 in the Population with CP were analyzed. Statistical methods included Analysis of Variance (ANOVA), Analysis of Covariance (ANCOVA), Linear Trend Test, and Standard Error Assessment.

**RESULTS:** A statistically significant difference exists between PODCI subscales in General Population and Population with CP across age groups and GMFCS levels. Motor scales and Global Functioning increase with age in both populations and are inversely proportional to GMFCS level in the Population with CP. HRQOL measures decrease with age in both populations with Happiness decreasing more in the General Population than those with CP as age increases.

**CONCLUSIONS:** PODCI demonstrates a statistically significant difference in motor performance and HRQOL in children ages 2-18, between the General Population and the population with CP. PODCI is a valid performance assessment tool for use in CP ages 2-18 across all GMFCS levels.

## INTRODUCTION

In 2005 we completed a multicenter study comparing assessments in the different domains of the International Classification of Functioning, Disability and Health (ICF) to correlate assessments in the clinic setting and <u>performance</u> in the community, comparing findings with a well-documented, clearly established measure of participation/performance for children of school-age, the School Function Assessment (SFA) [1], used as criterion for comparison. Measures used were Gross Motor Function Measure (GMFM), Gillette Functional Assessment Questionnaire (GFAQ), Three-dimensional Motion Analysis Gait velocity, Pediatric Quality of Life Inventory Generic Core Scales (PedsQL), and PODCI. Two articles have been published with some findings from this work [2, 3]. There is much more statistical evaluation from that study, yet to be published (personal communication). Of the measures assessed, PODCI and PedsQL had mild to strong correlations with all scales of SFA; PODCI had the greatest number of medium to strong effect sizes. Both PODCI and PedsQL use a transdiagnostic approach, Dalgleish et al [4], mirroring the World Health Organization (WHO) thrust in the International Classification of Functioning, Disability, and Health (ICF). Note we focus in this report on Performance and HRQOL.

Gates et al [2] validated PODCI as a pediatric performance tool for children with CP in ambulatory children from the clinical multicenter study, directly correlating PODCI with the SFA, which evaluates a child’s ability to participate in academic and social functioning in the school setting. The SFA is the ultimate measure of a patient’s school performance based on function. Capacity to walk 50 feet does not necessarily mean that they function well at school or work. PODCI had good correlation with the SFA, ecological validity, thus being chosen as an outpatient assessment tool to provide the accurate assessment of Performance in a clinic setting.

PODCI [5,6,7] evaluates children with disorders having an impact on the musculoskeletal system, to collect patient- and proxy-reported performance in multiple subscales including Motor scales, Health Related Quality of Life (HRQOL) and Comorbidity scales. The focus in this comparison is proxy report. In clinical practice, PODCI is used effectively to assess effectiveness of treatment regimens, and in research, to study performance outcomes [8,9,10]. PODCI is public domain, widely available, with a general population reference (online supplements – PODCI form and general population database) not available for condition-specific measures and allows transdiagnostic assessment.

Our literature review revealed 208 articles using PODCI (some early articles do not show under the name PODCI because the name had not yet been standardized), including continued strong usage of PODCI despite numerous other instruments being available.

**Figure 1.**
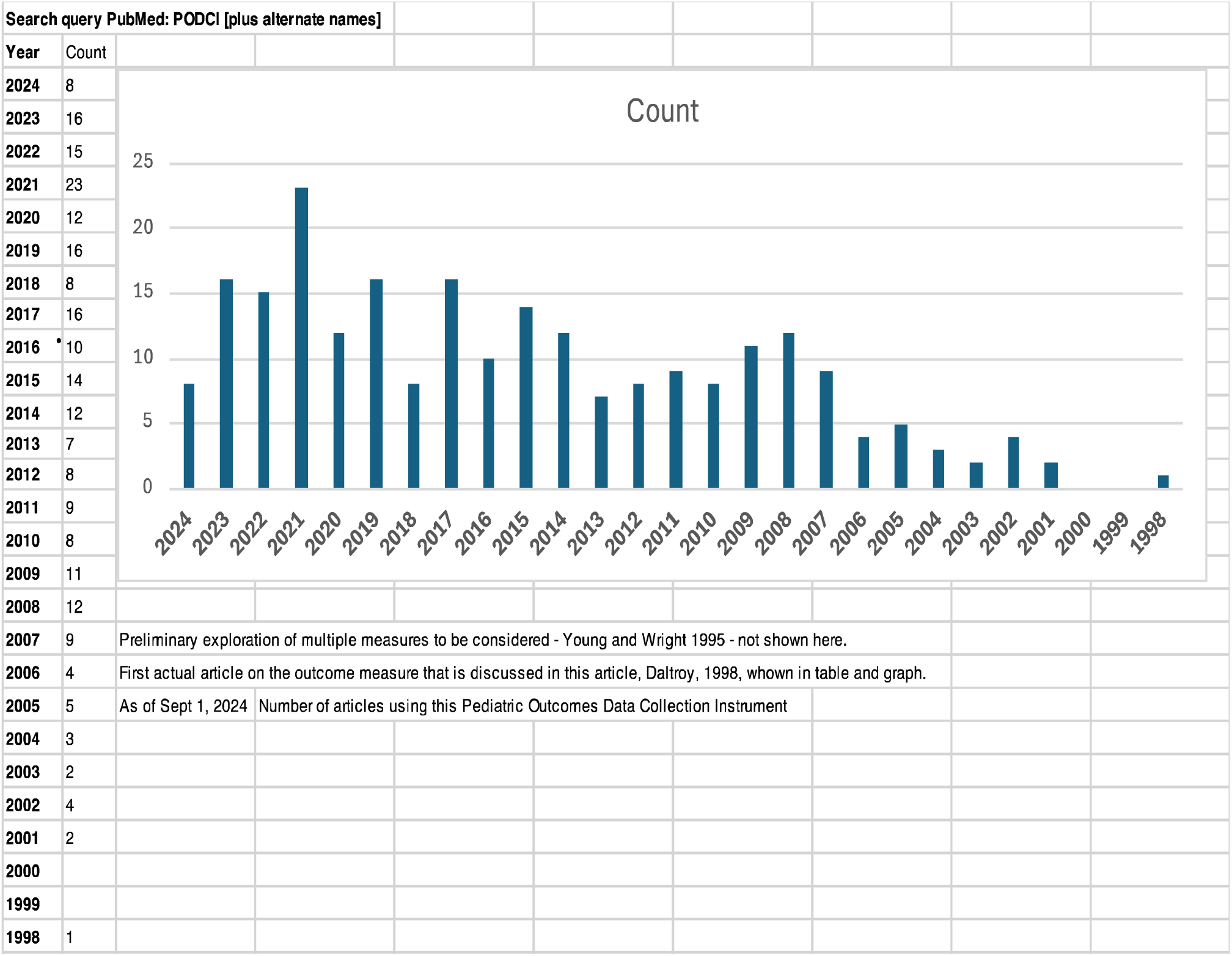
PubMed Articles Using PODCI.

CP is a clinically described disorder of neuromuscular development diagnosed in early stages of life, with a wide range of severity. It is one of the most common forms of developmental disability, affecting from 1 to nearly 4 per thousand children in the United States [11]. CP is the most frequently assessed condition with musculoskeletal impact using PODCI. Our literature review of 208 articles using PODCI showed 30.7% (n=64), focused on CP. Brachial Plexus Birth Palsy was second most frequently assessed condition using PODCI with 6.3% (n=13) of the 208 articles. PODCI is used in a wide variety of disorders. The literature review showed minimal comparison with the typically developing population, and in CP included primarily GMFCS levels I-III, with minimal representation of GMFCS IV, none in GMFCS V.

GMFCS is a five-level performance classification system, not an outcomes measure, based on level of functional mobility in patients with CP. Levels I is least functionally limited and level V most functionally limited [12,13,14]. GMFCS stability over time, regardless of treatment, has been reported from 72% to 84%-89% [15,16], though function changes with age. Currently available interventions are not expected to improve GMFCS levels but should at least aim to make meaningful functional improvements within a GMFCS level. PODCI affords assessment of such functional improvements on a finer gradational scale.

Hunsaker et al [17] indicated the need for a comparative group to help clinicians demonstrate positive outcomes of treatment. Hunsaker’s group established a national orthopaedic normative outcomes database using PODCI, delineating children in the General Population of the United States by age, sex, and comorbidities. The data showed statistically significant findings among demographic categories on PODCI subscales in the General Population [18]. We aim to extrapolate these findings to the Population with CP across GMFCS levels and age groups, with the General Population data as a comparative group.

In addition to a comparative group, baseline assessments are an important first step before assessing pre/post treatment outcomes. Kendall et al emphasized the importance of baseline assessments for measuring outcomes of treatment [19]. Use of PODCI as a baseline assessment in individual patients and as a standardized measure across age groups and GMFCS levels is the first step. As GMFCS motor development curves have been used relative to prognosis [20], correlation with PODCI in longitudinal studies, as a next step, may provide curves of anticipated function and may help with plan of care, prognosis, and management of expectations.

We stress looking at Performance transdiagnostically based upon the WHO thrust. We focus attention on this common language of performance in daily life, to help clinicians and researchers alike to be very comfortable using PODCI as a measure of functional performance. This does not argue against the use of other assessments: capacity/capability, body structure and function, or genetic testing. Our goals in managing patients should be based what they can do in everyday life [21], the purpose of “Functioning, Disability and Health, “in ICF. The intent of this paper is to demonstrate that PODCI differentiates between the General Population and the population with CP, in all GMFCS levels in children 2-18 years. Establishing statistically significant findings using PODCI in all GMFCS levels in children with CP is a first step in use of PODCI in future performance outcomes assessment in all levels of severity.

## METHODS

### General Methods

In 2000, an IRB-approved (Louisiana State University, Shreveport) database of children with CP was established due to a paucity of CP registry data in the United States. The sample was a convenience sample from pediatric orthopaedic referral hospitals, the greatest number from a six-state area in the Southern United States but including children from 14 states. Data was collected between 2000 and 2020. The questionnaire was made available to all caregivers bringing a child with CP to outpatient clinic or motion analysis center appointments and completed on paper during the visit. The validated Spanish version was used for Spanish-speakers [22]. The database includes 2470 children, ages 2-18 years, with CP presenting to the clinic or motion analysis center for evaluation. Pediatric orthopedic providers experienced in neuromuscular pathologies assigned GMFCS levels to children with CP according to published criteria [14 CanChild GMFCS-ER]. Cross-sectional data obtained from PODCI during the first assessment of each child at our institutions serves as a baseline measure. The PODCI Questionnaire was completed by parents/caregivers regarding the child’s performance and contextual factors. Only questionnaires completed by proxy are included to maintain consistency with the methods used for the General Population database, avoiding “cross-informant variance.” The General Population database established by Hunsaker et al [17] used single wave mailing methodology including 5238 proxy responses to PODCI in the pediatric population in the United States. This data comprises the comparative database used in the current study.

The age range in both populations was from 2-18. The mean age of all children combined is 10.94 (SD 4.58). The mean age of children with CP is 9.38 (SD 4.40); the mean age of the children in the General Population is 11.65 (SD 4.48), thus those with CP are somewhat younger.

PODCI is a performance measure (qualifier for Participation Scale of ICF) [21 ICF Beginners Guide] as opposed to a capability/capacity measure. It is an assessment of the child’s daily life. PODCI consists of the following subscales: Upper Extremity and Physical Function, Transfers and Basic Mobility Tasks, Sports and Physical Functioning (higher level functioning), Pain Free/Comfort, and Happiness. The Global Functioning Scale consists of the mean of the first four scales (Upper Extremity, Transfers and Mobility, Sports and Physical Functioning, and Pain Free/Comfort). PODCI also includes expectations of treatment scale and a satisfaction with treatment scale, not used in this cross-sectional study. (See online supplement for PODCI form) Demographic and Comorbidity data are also collected.

### Statistical Methods

Means and standard errors were used to summarize PODCI values by age. Since distribution of numbers of participants in GMFCS levels in the population with CP were not consistent across age levels, adjusted means for PODCI values by age rather than raw mean values were used for those with CP. See Table 1. This data indicates there were more severely-involved children seen in the youngest age groups with fewer severely-involved children in the older age groups. Since the clinics are referral centers with large geographical bases, the patients enrolled were those referred. There was no systematic selection bias at our hospitals; all patients with CP were approached to participate. Attempts to explain the variations would be purely speculative. (See comment in Limitations section.)

**Table 1:** Patient Age by GMFCS Level (Frequency and Percentage)

| Age Category | GMFCS Level |  |  |  |  |  |
| --- | --- | --- | --- | --- | --- | --- |
| Frequency | I | II | III | IV | V | Total |
| 2-6 Years | 126 | 249 | 133 | 137 | 73 | 718 |
| 7-13 Years | 241 | 469 | 248 | 149 | 94 | 1201 |
| 14-18 Years | 112 | 200 | 109 | 52 | 36 | 509 |
| **Total** | 479 | 918 | 490 | 338 | 203 | 2428 |
| Age Category | GMFCS Level |  |  |  |  |  |
| Percentage | I | II | III | IV | V | Total |
| 2-6 Years | 17.55 | 34.68 | 18.52 | 19.08 | 10.17 | 100 |
| 7-13 Years | 20.07 | 39.05 | 20.65 | 12.41 | 7.83 | 100 |
| 14-18 Years | 22 | 39.29 | 21.41 | 10.22 | 7.07 | 100 |
| **Total** | 19.73 | 37.81 | 20.18 | 13.92 | 8.36 | 100 |

Because the distribution is skewed, with more severe children in the younger age group, and fewer in the older age group, adjusted means are used.

Adjusted means for unequal GMFCS values across ages present estimates of anticipated raw mean values in the children with CP as if GMFCS levels were equivalent at each age (an accepted statistical technique). Adjusted mean values were computed for children with CP using an Analysis of Covariance (ANCOVA) model where the PODCI value was the dependent variable and GMFCS level and age were independent variables.

A one-way Analysis of Variance (ANOVA) model was used to compare PODCI means in the two populations, General vs CP, at each age, where the PODCI value was the dependent variable and population was the independent variable. A one-way ANOVA to compare PODCI means among GMFCS levels was used to determine if PODCI values were significantly different across GMFCS levels. If this initial F-test was statistically significant, a linear trend contrast of the PODCI means was performed to determine if this statistically significant difference showed a linear trend.

All statistical tests were two-tailed and conducted at the 0.05 significance level using SAS® Version 9.4.

## RESULTS

There were no statistically significant differences between male vs female on PODCI. In both the General Population and the Population with CP, there were more males represented than females, 51.48% to 48.52% M to F in General Population: 55.39% to 44.61% in Population with CP. Table 2 shows Spearman correlations between GMFCS Level and PODCI Scale, with effect sizes [23 Cohen]

**Table 2:** Spearman Correlations between GMFCS Level and PODCI Scales.

| PODCI Domain | Spearman Correlation Coefficient | Effect Size |
| --- | --- | --- |
| Upper Extremity and Physical Function | -0.622 | Large Negative |
| Transfers and Basic Mobility | -0.777 | Large Negative |
| Sports and Physical Function | -0.729 | Large Negative |
| Pain Free / Comfort | -0.074 | No Effect |
| Happiness | -0.156 | Small Negative |
| Global | -0.719 | Large Negative |

Because GMFCS levels are discrete ordinal variables and PODCI scales are continuous variables, Spearman Correlations are used. Effect Size is based upon Cohen [23]. Negative effect size due to worsening GMFCS with higher level, improving PODCI value with higher level.

ANCOVA and linear trends displayed a statistically significant difference (linear trend p < 0.0001) between the General and CP Populations in all PODCI subscales when compared by age. Subscales measuring motor functioning increased in both populations as age increased, and for HRQOL scales of Happiness and Pain Free/Comfort, decreased in both populations as age increased, However, the magnitude of change was greater in the General Population compared to those with CP (Figures 2 and 3).

**Figure 2.**
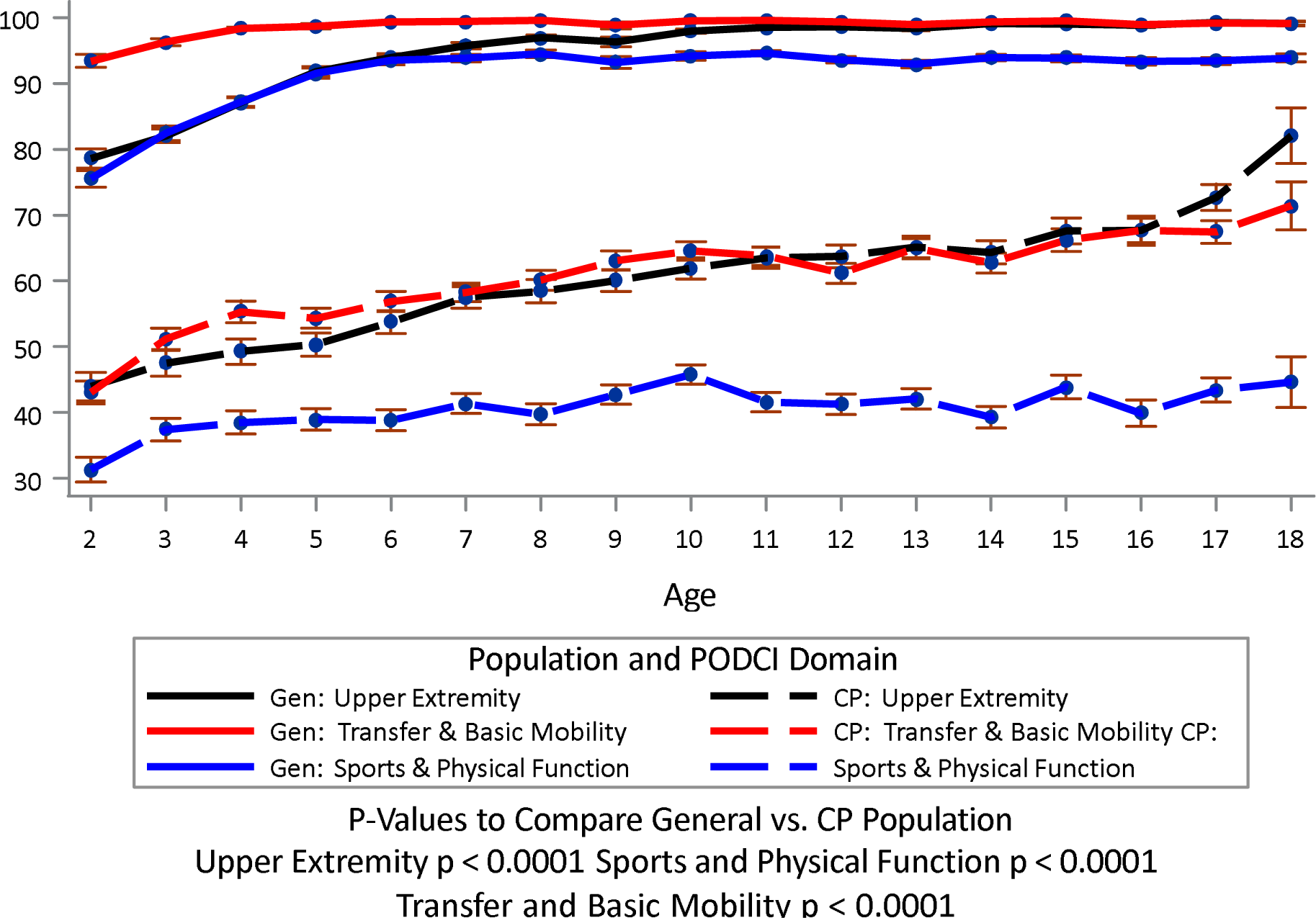
PODCI Motor Function Means and Standard Errors by Age General vs. Cerebral Palsy Populations.

**Figure 3.**
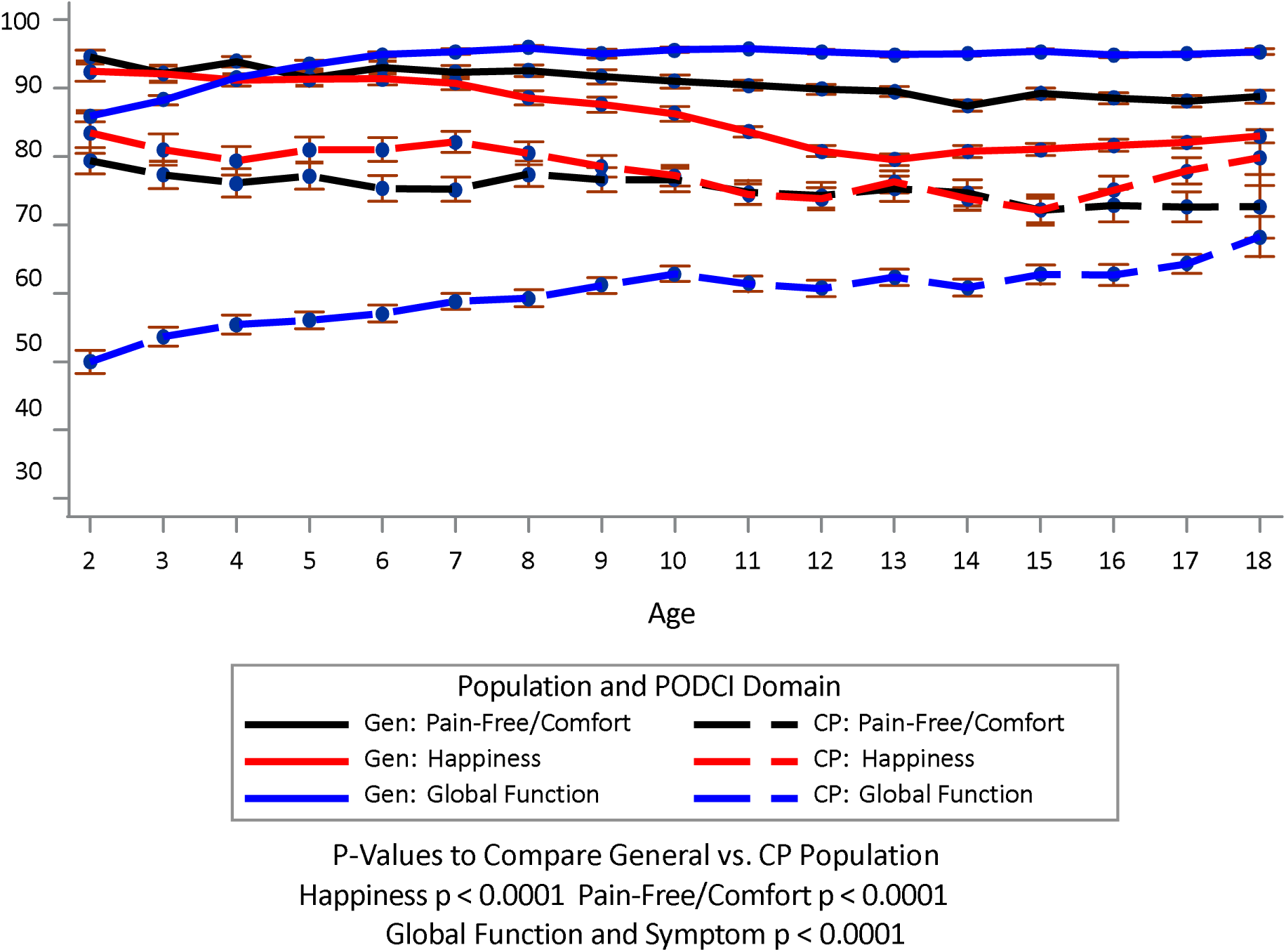
PODCI Perceived Well-Being Means and Standard Errors by Age General vs. Cerebral Palsy Populations.

One-way ANOVA tests demonstrated a clear difference between PODCI scale means in the General and CP Populations across GMFCS levels I-V. In each PODCI subscale a statistically significant negative linear trend exists (p < 0.0001) in PODCI means from the General Population to each of the five GMFCS levels for the Population with CP (Figures 4-9). A negative linear relationship is seen.

**Figure 4.**
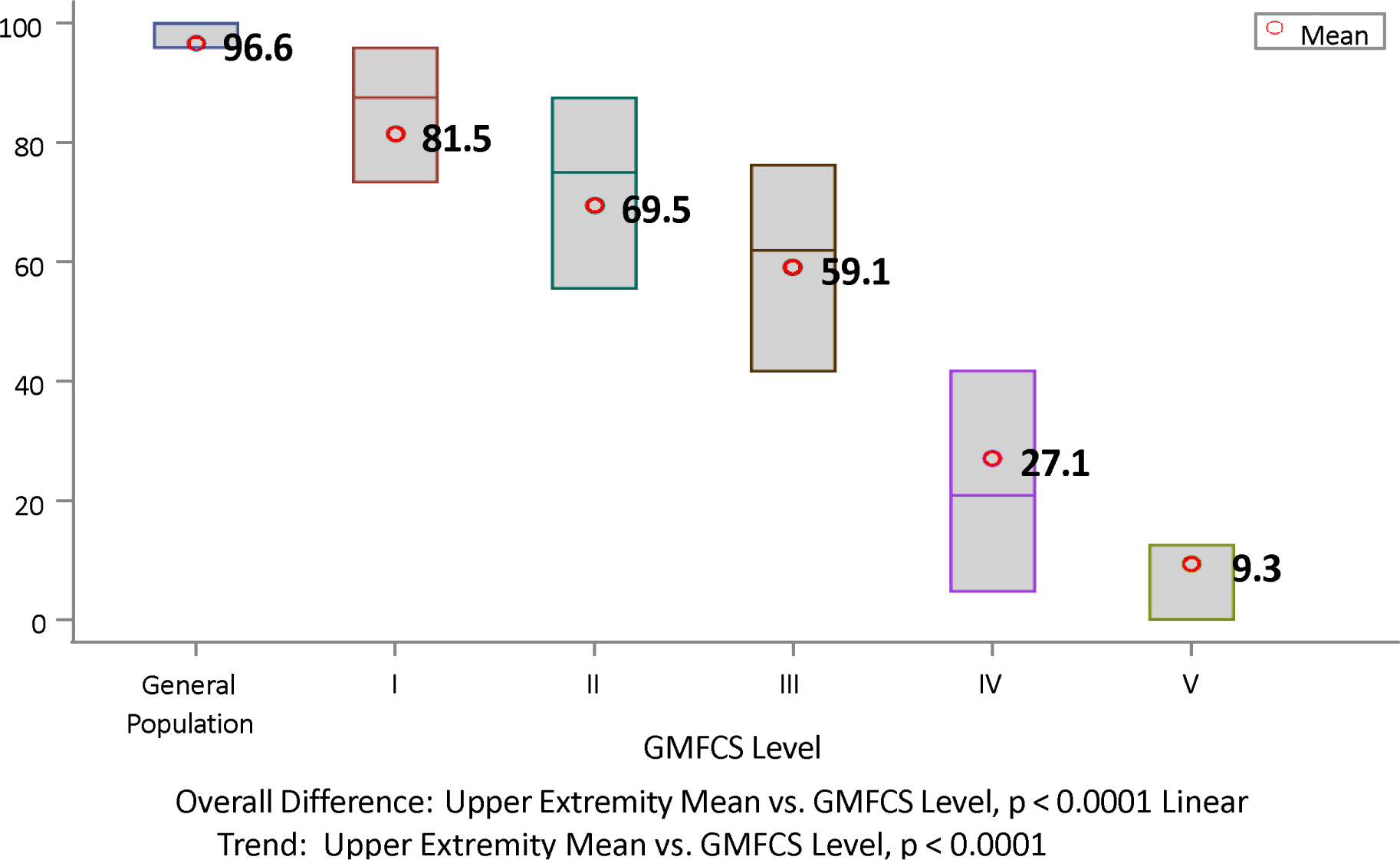
Upper Extremity Means by GMFCS Level.

**Figure 5.**
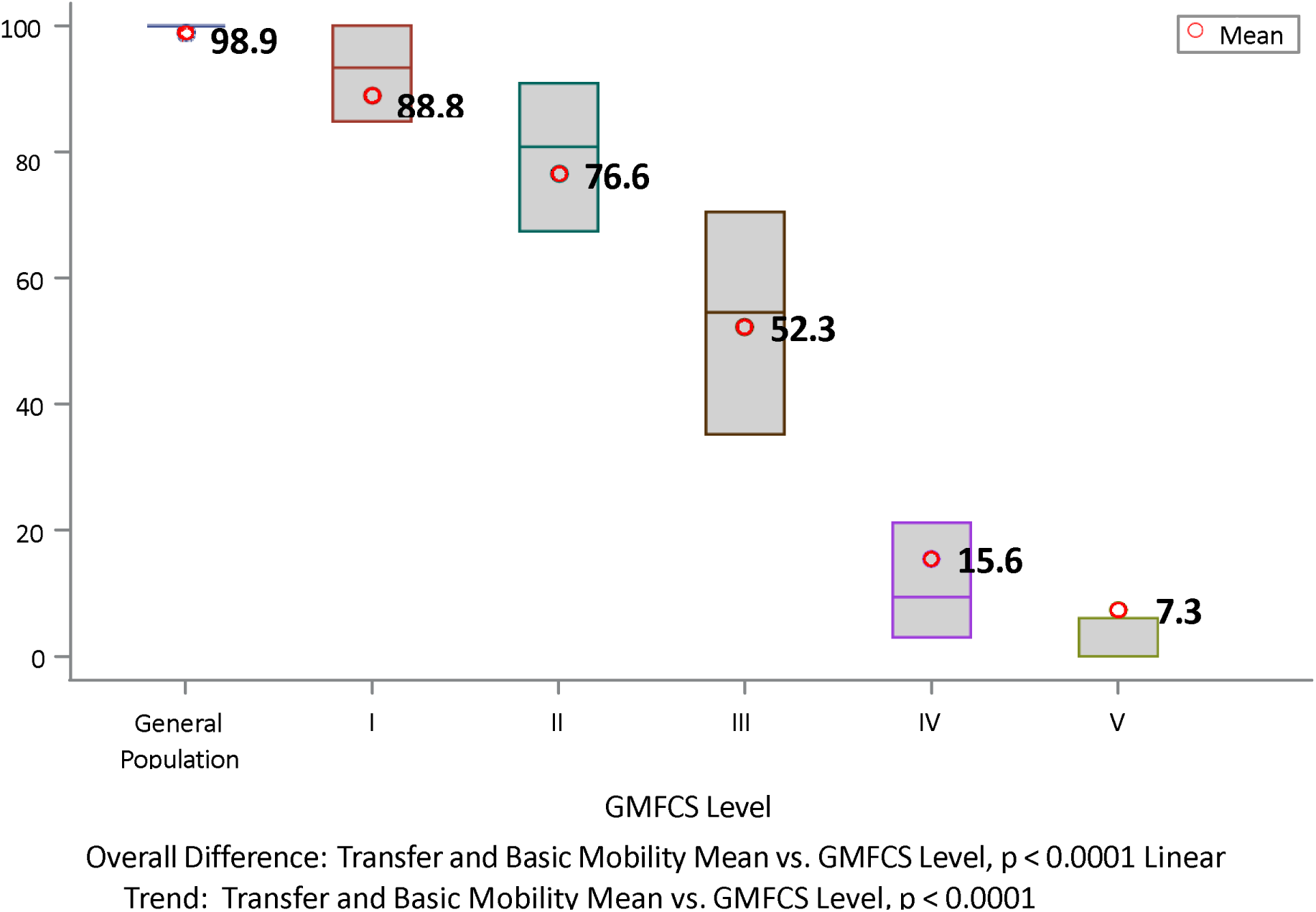
Transfer and Basic Mobility Means by GMFCS Level.

**Figure 6.**
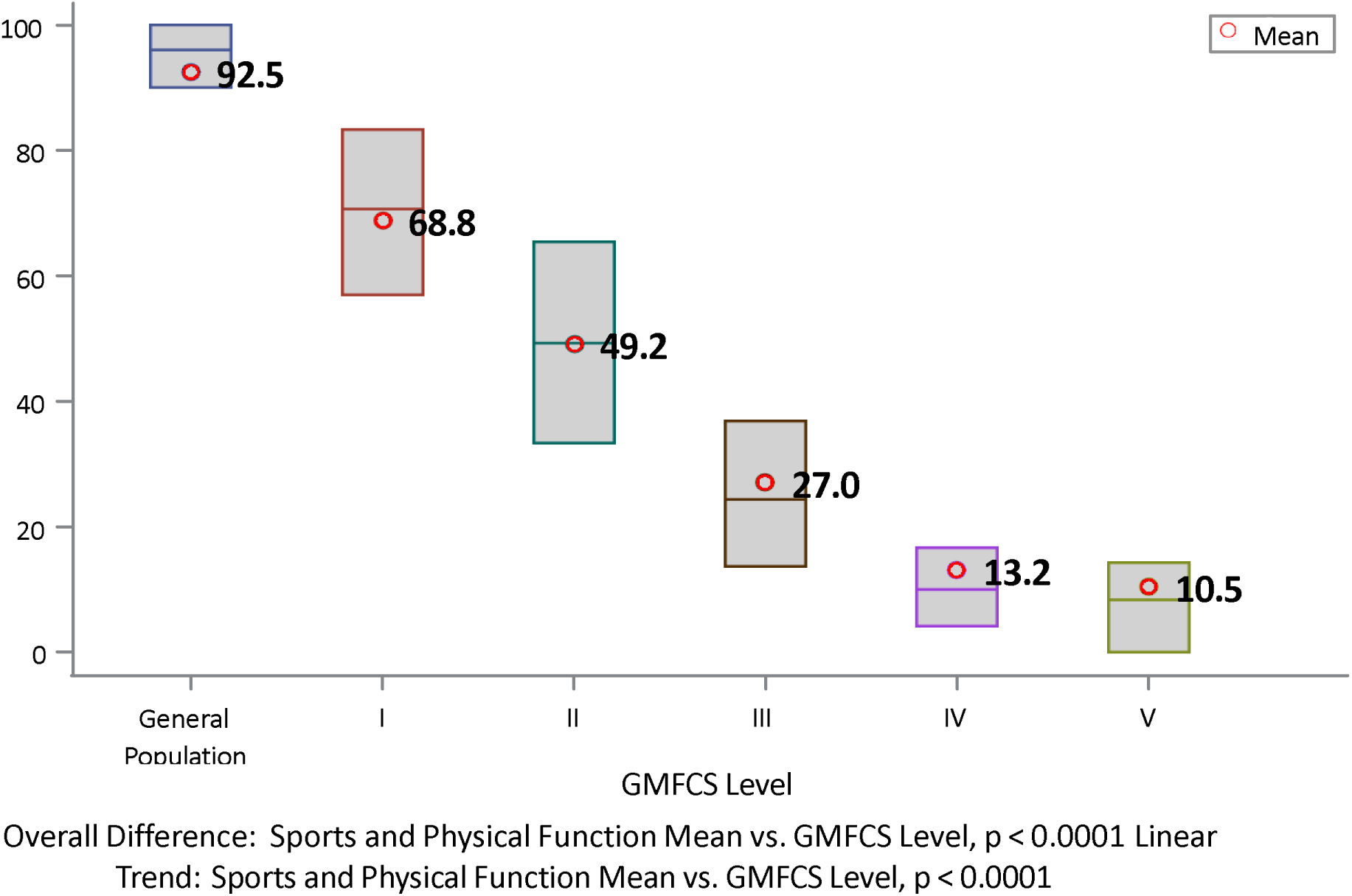
Sports and Physical Function Means by GMFCS Level.

**Figure 7.**
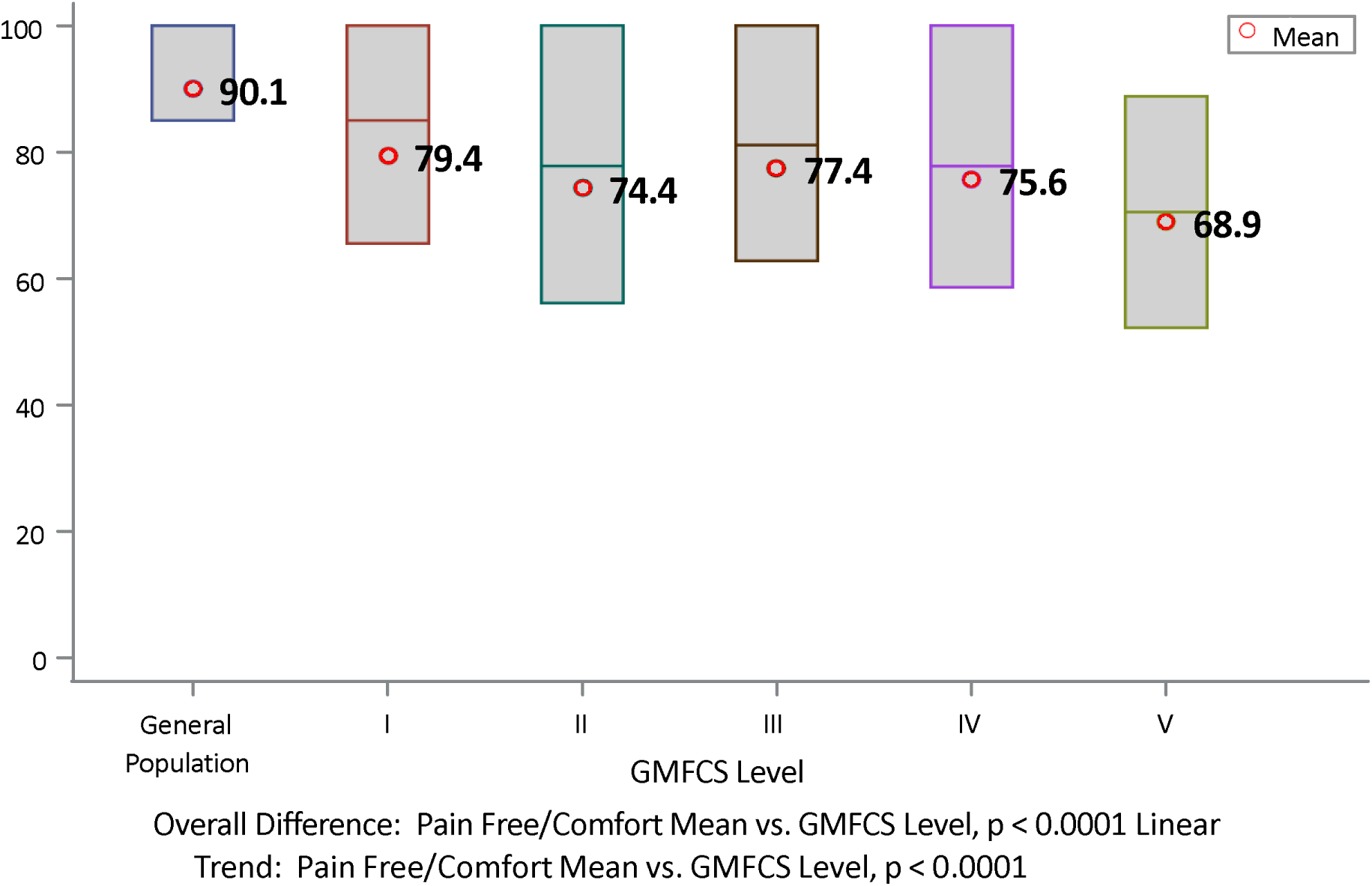
Pain Free/Comfort Means by GMFCS Level.

**Figure 8.**
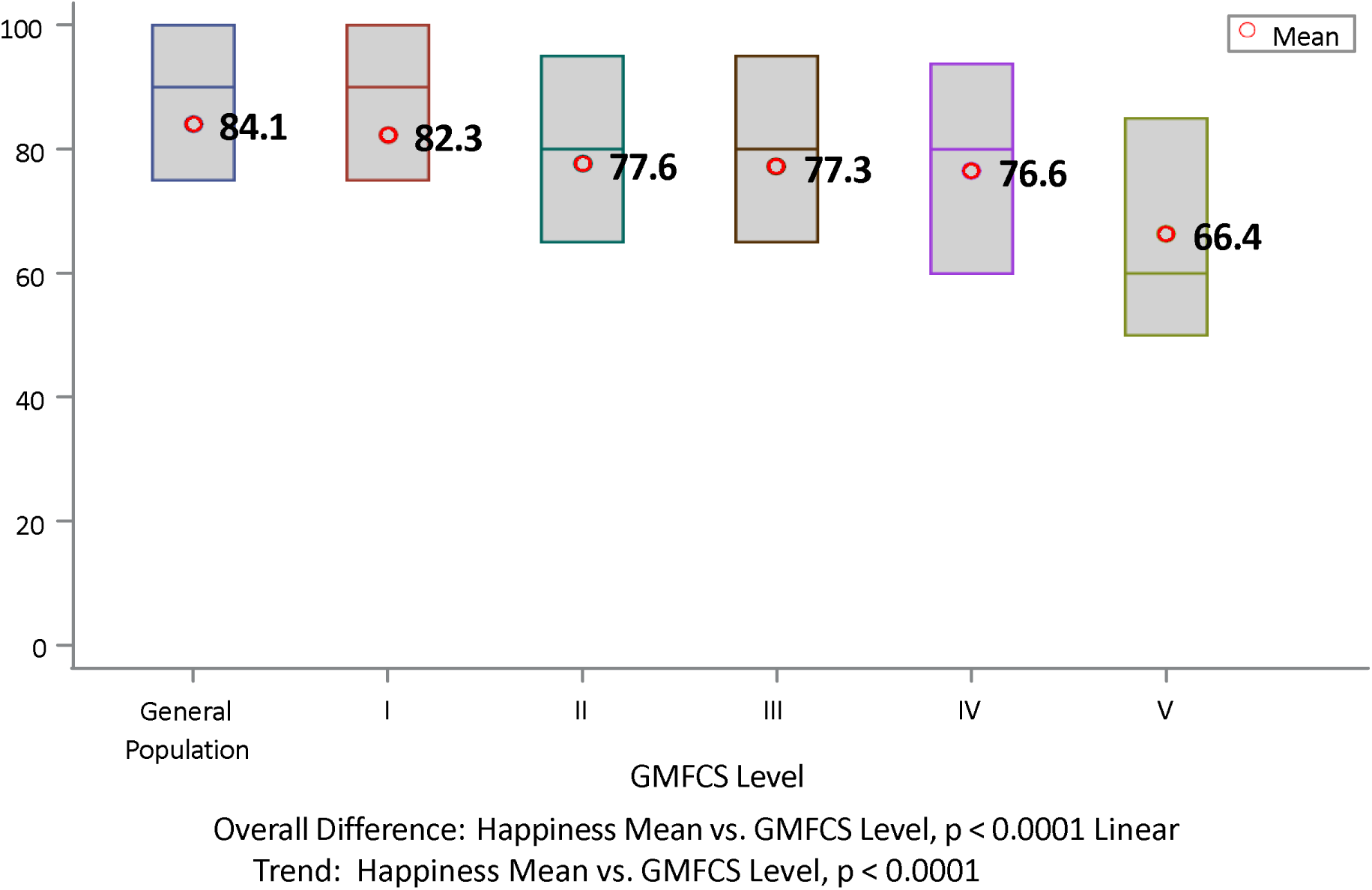
Happiness Means by GMFCS Level.

**Figure 9.**
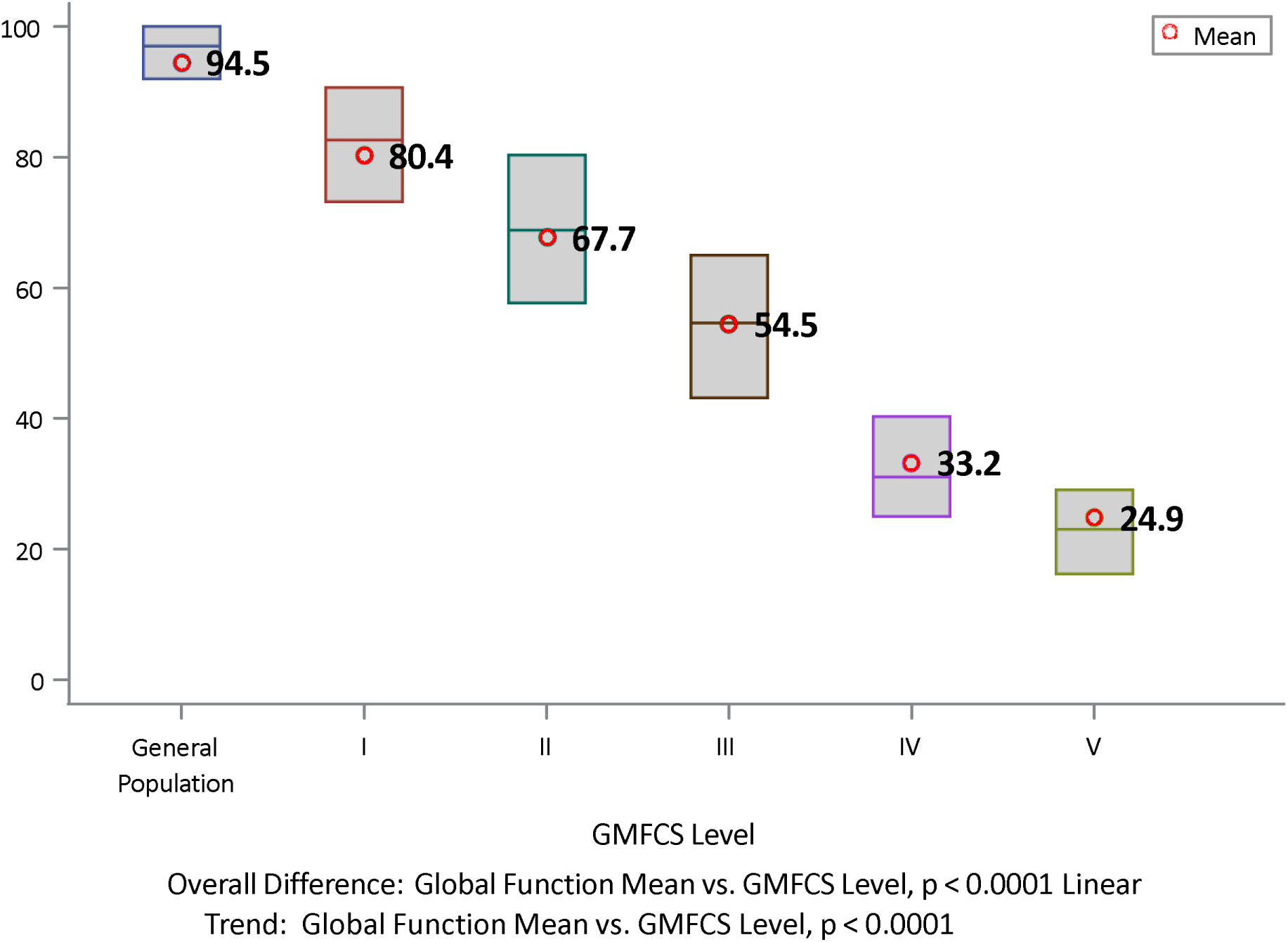
Global Function Means by GMFCS Level.

**Figures 4-9** show each individual PODCI scale, comparing the General Population to each of the five GMFCS levels in the CP Population. p values show the significance of the differences across each PODCI scale domain within the population groupings. All linear trends are p<0.0001.

## Discussion

We have focused attention on the common language of functional performance in daily life, to help clinicians and researchers alike be comfortable using PODCI as a performance assessment tool, in line with the ICF and transdiagnostic approach. The goal was to demonstrate that PODCI differentiates between General and CP Populations and among GMFCS functional levels, in all GMFCS levels in children 2-18 years. The PODCI Questionnaire was shown to differentiate between children in General and CP Populations at all severity levels of GMFCS and across age groups.

The pattern as children age, in this cross-sectional sample, is that motor and global functioning increase with age. This trend is demonstrated by PODCI in both General and CP Populations, although a more vertical increase in motor scales of TBM, UE, SPF, and Global Functioning exists in the General Population compared to those with CP. Children with CP show a more modest increase in these scales but still demonstrate a positive slope with age (<u>new information</u>). Anecdotal reports suggest decreased function with age.

Since GMFCS levels are an empirically-created scale constructed around daily functional mobility (motor performance – [21 ICF Beginners Guide], shown to correlate with GMFM, it would be expected for tasks related to mobility and purposeful movement to be increasingly impaired as GMFCS level increases [24,25]. Hence, PODCI subscales were inversely proportional with GMFCS level, demonstrating decreasing performance as level of severity of CP increased.

PODCI HRQOL scales of Happiness and Pain Free/Comfort demonstrated a divergence between General and CP Populations with respect to age. There was an inversely proportional relationship between age and Happiness and Pain Free/Comfort in both populations. However, the General Population showed a marked decrease in these scales with increasing age compared to the Population with CP, which demonstrated a more modest effect and remained elevated across age groups. There is a less significant effect on Happiness and Pain Free/Comforrt in children with CP as level of severity of disease increases, indicating that reported happiness and comfort levels are better than expected in children with more severe motor involvement. (See comment in Limitations section.) Such information helps clinicians remember that treatment for improving motor functioning does not necessarily quantify the same benefit in HRQOL scales as it does in the motor functioning scales. These findings should be considered when discussing treatment options with patients and families to provide a holistic analysis of specific interventions.

Though GMFCS is not an outcomes measure and improvement in GMFCS level is not an intended outcome of treatment, PODCI scales allow a finer gradation of function within GMFCS levels. Outcomes on PODCI have been shown to improve with surgical treatment [26], even to the point of discrimination between surgical interventions. The current baseline data should serve as a basis upon which to evaluate future interventional studies.

## CONCLUSIONS

CP is the most common pediatric neuromuscular pathology, affecting 1 per 345 children in the United States [8]. Assessment tools including the PODCI Questionnaire have been used as performance/outcomes measures for individuals in the General Population and for children with disorders affecting the musculoskeletal system. PODCI has been used extensively to assess motor performance and HRQOL in CP. The current study expands assessments to include all GMFCS levels and all ages 2-18. Additionally, no PODCI correlations have been established between General and CP Populations.

This study demonstrates PODCI is an effective tool for clinicians to evaluate performance levels and HRQOL. This is only a first step toward use in outcomes assessment in CP. Utilization of PODCI for individual treatment plans for patients with CP, with comparative pre- and post-intervention assessments (future work) may help predict clinical outcomes, guide individual plans of care, and manage expectations of outcomes. Future studies, including longitudinal research, comparing additional demographics across GMFCS levels using contextual and personal factors are needed to establish use for treatment outcomes and guide clinical practice. There is no intent to supplant condition-specific assessment tools in children with cerebral palsy, but to provide a performance assessment tool that can be reliably used among children with childhood-onset disabilities transdiagnostically, in an era of increasingly compartmentalized diagnosis and management.

### Limitations

Limitations to this study include the convenience sample of the patients with CP, the geographic demographics of the population, the cross-sectional data analysis, and the variability of ages within the GMFCS levels. The data from the children with CP was created from a convenience sample of parents/caregivers presenting with their affected child for outpatient clinic and/or motion analysis visits. The demographics for the data collected included 14 states but may not be representative of the entire pediatric population with CP, since the greatest number were referred from six states in the mostly rural, southern United States. A longitudinal study including data from a smaller sub-population in the CP data set is currently underway to support the findings of this cross-sectional data set. Lastly, variability in numbers of children of different ages within GMFCS levels required the calculation of adjusted means as opposed to the use of raw data. Two studies have reported some variation in rates of children being reclassified into different GMFCS level (i.e., GMFCS level is not entirely stable over time), affecting the youngest age groups [15,25]. (This <u>could</u> be the answer to our variation with age.)

Some may have concerns that proxy reports are used rather than patient-reported outcomes. The rationale was thoroughly discussed above, but there is much more detailed discussion about comparing self-versus-parent report with PODCI in the Population with CP in an article in Developmental Medicine and Child Neurology 2010 [27]. Two comments from that article pertinent here were that the overall pattern was one of greater concordance in more severely affected adolescents, and when there was discrepancy, adolescents with CP reported themselves higher than parents, which would, therefore, suggest even greater functional gains and HRQOL with age, which would further strengthen our findings. When possible, practitioners should seek input from patients and parents and consider each in their own right. Within the confines of assuring comparability of data in this report, on all participants, all ages, all GMFCS levels, and comparing with the General Population, parent report was the only choice, to avoid “cross-informant variance,” not comparing “apples to oranges.”

## Supporting information

General Population Raw data

## Data Availability

The General Population database is available and included in the submission but data from our site's population will not be shared.

